# Sociodemographic inequity in COVID-19 vaccine uptake among youth in Zimbabwe

**DOI:** 10.1101/2023.03.10.23287107

**Authors:** Leyla Larsson, Chido Dziva Chikwari, Victoria Simms, Mandikudza Tembo, Agnes Mahomva, Owen Mugurungi, Richard Hayes, Constance Mackworth-Young, Sarah Bernays, Constancia Mavodza, Tinotenda Taruvinga, Tsitsi Bandason, Ethel Dauya, Rashida A Ferrand, Katharina Kranzer

## Abstract

**Introduction:** COVID-19 vaccine acceptance research has mostly originated from high-income countries and reasons why youth may not get vaccinated may differ in low-income settings. Understanding vaccination coverage across different population groups and the sociocultural influences in healthcare delivery is important to inform targeted vaccination campaigns.

**Methods:** A population-based survey was conducted in 24 communities across three provinces (Harare, Bulawayo and Mashonaland East) in Zimbabwe between October 2021 and June 2022. Youth aged 18 - 24 years were recruited using random sampling. Data on sociodemographic information and COVID-19 vaccination uptake and reasons for non-uptake were collected.

**Results:** A total of 17,682 youth were recruited (n=10,743, 60.8% female). The median age of survey participants was 20 (IQR: 19 – 22) years. Almost two thirds (n=10,651, 60.2%) of participants reported receiving at least one dose of COVID-19 vaccine. A higher proportion of men than women had been vaccinated (68.9% vs 54.7%), and vaccination prevalence increased with age (<19 years: 57.5%, 20-22: 61.5%, >23: 62.2%). Lack of time to get vaccinated, belief that the vaccine was unsafe and anxiety about side effects (particularly infertility) were the main reasons for not getting vaccinated. Factors associated with vaccination were male sex (OR=1.69, 95%CI:1.58-1.80), increasing age (>22 years: OR=1.12, 95%CI:1.04-1.21), education level (post-secondary: OR=4.34, 95%CI:3.27-5.76), and socioeconomic status (least poor: OR=1.32, 95%CI:1.20-1.47).

**Conclusion:** This study found vaccine inequity across age, sex, educational attainment and socioeconomic status among youth. Strategies should address these inequities by understanding concerns and tailoring vaccine campaigns to specific groups.

**What is already known on this topic:** Many countries have faced challenges when rolling out COVID-19 vaccines. Infrastructure, logistics, misinformation and vaccine hesitancy have been barriers to vaccine access and uptake globally. Vaccine nationalism by high-income countries has particularly affected countries in Africa and Asia, resulting in inequity between countries and regions.

**What this study adds:** Vaccine uptake among youth in Zimbabwe was more than 50% across all age-groups. Men, those with more education and those living under less socially deprived socioeconomic conditions were more like to be vaccinated. Fear of side effects and myths circulating on social media were barriers. Religion was less of a barrier than other studies reported, likely due to religious institutions’ collaborations in COVID-19 vaccination efforts.

**How this study might affect research, practice or policy:** Vaccination campaigns should actively address specific concerns of communities, especially concerns around fertility and early death, and provide vaccines in easy-access and convenient locations. Involving community leaders in both education and vaccination efforts is pivotal given the trust and influence they have.

## Introduction

By February 2023, more than 750 million SARS-CoV2 infections and 6.8 million COVID-19 associated deaths had been reported globally [1]. The development of vaccines against SARS-CoV2, which primarily protect against severe disease, has however greatly reduced both COVID-19-related mortality and morbidity [2,3]. Global vaccination programmes were fast-tracked under the COVID-19 Vaccines Global Access (COVAX) initiative at the beginning of 2021 [4]. Vaccination rates have, however, failed to meet the targets of vaccinating 70% of the world’s population against COVID-19 by mid-2022 set by the World Health Organization (WHO), especially in the African continent [5].

By February 3^rd^ 2023, Africa has reported a vaccination coverage (receiving at least 1 dose of a COVID-19 vaccine) of eligible population of 46% and of total population of 27%, with coverage varying greatly from < 6% in Madagascar, Cameroon and the Democratic Republic of Congo to more than two thirds of the eligible population in Rwanda and Liberia [6,7]. Differences are partly explained by availability of vaccines, infrastructural constraints, and COVID-19 vaccine nationalism in high income countries (HIC) leading to insufficient donations of vaccine to low- and middle-income countries (LMICs) [8]. In addition, vaccine hesitancy has been reported to be one of the main barriers towards meeting global vaccine coverage targets [9-11]. Studies conducted both in HICs and LMICs have reported widespread hesitancy towards receiving the COVID-19 vaccine, despite most of the population having previously received vaccines for other viruses [12,13] Zimbabwe recorded its first case of COVID-19 in March 2020 and was subsequently among the first African countries to implement vaccination, with the first vaccine administered on 18 February 2021 [14]. In contrast other countries in the region, Zimbabwe procured COVID-19 vaccines (SinoPharm and SinoVac) through a bilateral agreement with China [15,16]. While non-availability of vaccines was a major barrier for vaccine uptake in other African countries, this was not the case in Zimbabwe [16]. Initially, healthcare workers and people working at borders were targeted for COVID-19 vaccination, followed by those with chronic conditions and essential workers such as teachers [17]. With the availability of more vaccine doses, eligibility was rapidly extended to the rest of the adult population (people aged 18 and over). In November 2021, vaccines were also made available to adolescents aged 16-18 years and in March 2022 to children aged 12 years and older [16-18]. The initial vaccine schedule was two doses of either SinoPharm or SinoVac three weeks apart. A third booster dose was introduced in January 2022 [19-21].

The Zimbabwe vaccination campaign was administered through vaccination centres established in hospitals and clinics and outreach services. In addition, the vaccination campaign included educational programmes in schools, national mobilisation of frontline workers to assist in getting to hard-to-reach populations, and nationwide health education broadcasting [18,22-23]. Once vaccination eligibility was extended to adolescents the government partnered with organisations such as United Nations Children’s Fund (UNICEF) to generate vaccination messaging taregting children and adolescents [18,25-27]. As of February 3^rd^ 2023, the Zimbabwean Government estimates that 44% of the total population have received at least one COVID-19 vaccine dose. With approximately 60% of the population being eligible (12* years), coverage among those eligible was 77% [1,2].

Despite the tremendous efforts put into the vaccination campaign, a considerable proportion of the general population remains unvaccinated and more recently vaccination coverage has stagnated [14]. While a Partnership for Evidence-Based Response to COVID-19 survey in September 2021 demonstrated that 82% of respondents in Zimbabwe were satisfied with the government’s response, there was still some vaccine hesistancy [28]. Findings from a survey in Zimbabwe in January 2022 indicated that half of the population displayed some vaccine hesitancy, mostly driven by social media misinformation and lack of trust in official information [29]. Furthermore, there is a lack of evidence in the literature regarding uptake of COVID-19 vaccination among young people and potential causes of vaccine hesitancy in Zimbabwe.

This study investigates self-reported COVID-19 vaccine uptake among 17,862 young people aged 18-24 years across three provinces (Harare, Bulawayo, and Mashonaland East) in Zimbabwe and explores sociodemographic and -economic factors associated with uptake and reasons for possible vaccine hesitancy.

## Methods

### Enrolment

This study used data from a population-based survey which was conducted to ascertain the outcome of the CHIEDZA trial. CHIEDZA is a cluster randomised controlled trial conducted in three provinces (Harare, Bulawayo, and Mashonaland East) in Zimbabwe investigating the impact of providing community-based integrated HIV and sexual and reproductive health services to young people aged 16-24 years on population-level HIV outcomes (NCT03719521) [30]. Taking advantage of a large population-based survey being undertaken as the COVID-19 vaccination campaign was being rolled out, we sought to understand COVID-19 vaccination coverage among youth to inform future vaccination strategies. Youth were randomly selected using geographic information system (GIS) methods. The survey was conducted in Harare (October– December 2021), Bulawayo (January– March 2022) and Mashonaland East (April– June 2022)aiming to recruit 16,800 18-24 year-olds (5,600 per province). COVID-19 vaccines became eligible for youth (aged 18 and above) in March/April 2021 and for 16-18 year olds in November 2021 [17]. Sociodemographic data, self-reported COVID-19 vaccination, and reasons for not being vaccinated were collected using interviewer-administered questionnaires. Participants could select multiple reasons for non-vaccination from a predefined list, or give additional reasons which were recorded as free text.

### Data collection

Participants viewed an information video about the study on a tablet screen prior to providing electronic consent. Survey data were collected onto electronic tablets using SurveyCTO (Cambridge, USA). Data were stored on a server at the Biomedical Research and Training Institute (BRTI) in Zimbabwe and no identifying information was collected. This study was performed in accordance with the study protocol, the Declaration of Helsinki, as well as national and other regulatory guidelines.

### Data analysis

Data analysis was conducted using Stata v16.1 (StataCorp, USA). A descriptive analysis was performed using proportions for categorical data, medians and means for continuous data, followed by a univariable logistic regression analysis to investigate the association between sociodemographic characteristics (sex, age, educational attainment, socioeconomic status) and self-reported COVID-19 vaccine uptake. The outcome was self-report as having received at least one dose of a COVID-19 vaccine. Factors associated with the outcome in the univariable models were built into a multivariable logistic regression model, using only respondents with no missing data. To control for time, month of data collection was included in the model a priori. A Likelihood Ratio Test (LRT) was conducted for each variable to assess strength of evidence of association and reported as a p-value. The socioeconomic status variable was created using a principal component analysis of ownership of assets (refrigerator, bicycle, car, tv, radio, microwave, cell phone, and computer) and was then reported in quintiles. Analysis was performed for all provinces and then performed separately for each province as enrolment was conducted sequentially. Reasons for not taking up the vaccine were categorised according to the “5C’s” as described by Razai et al.: i) confidence (safety and efficacy of the vaccine), ii) complacency (perception of low risk and disease severity), iii) convenience (barriers and access), iv) communication (sources of information), and v) context (sociodemographic characteristics) [31].

### Ethics

The study received ethical approval from the Medical Research Council of Zimbabwe (MRCZ) (MRCZ/A/2387), the BRTI Institutional Review Board (IRB) (AP149/2018) and the London School of Hygiene and Tropical Medicine (LSHTM) ethics committee (16124).

### Patient and public involvement

The information video about the study was co-designed with and piloted among youth. The study questionnaire was also piloted with youth. The study had a Youth Advisory Board that provided guidance on study design and conduct. An extensive public engagement programme was undertaken alongside the study that included a national crowdsourcing competition to a) elicit young people’s perceptions about health issues in their communities and b) train youth as researchers through a mentored programme termed Youth Researchers Academy [32].

## Results

Of the 18,682 randomly sampled eligible youth in the study communities, 17,682 (94.6%) provided consent to participate. Of these, 5,849 (33.1%) were recruited in Harare, 5,969 (33.8%) in Bulawayo, and 5,864 (33.2%) in Mashonaland East. The median age of survey participants was 20 years (IQR: 19 – 22), and 10,742 (60.8%) were women. This higher proportion of women reflected the community composition.

Overall, 10,652 (60.3%) particpants self-reported having received at least one dose of a COVID-19 vaccine, with 8,316 (78.1%) having received two doses (Figure 1). The proportion vaccinated was higher among males (n=4,779, 68.9%) compared to females (n=5,872, 54.7%) and increased with age (<20: 57.5%, 20-22: 61.5%, >22: 62.2%) (Table 1). Overall, Harare province, where data collection preceded the other provinces by 3 (Bulawayo) and 6 (Mashonaland East) months, had the lowest proportion vaccinated (42.6%) compared to Bulawayo (69.6%) and Mashonaland East (68.3%) (Figure 1). The proportion vaccinated and double-vaccinated increased with each subsequent month during the survey in Harare and Bulawayo, but not in Mashonaland East.

**Table 1:** Sociodemographic characteristics and reported COVID-19 vaccination

|  | n | Vaccinated |
| --- | --- | --- |
| Sex |  |  |
| Male | 6,940 | 4,779 (68.9%) |
| Female | 10,742 | 5,872 (54.7%) |
| Age |  |  |
| < 20 | 6,809 | 3,916 (57.5%) |
| 20 – 22 | 4,702 | 2,894 (61.5%) |
| > 22 | 6,171 | 3,841 (62.2%) |
| Province |  |  |
| Harare | 5,849 | 2,492 (42.6%) |
| Bulawayo | 5,969 | 4,155 (69.6%) |
| Mashonaland East | 5,864 | 4,004 (68.3%) |
| Education |  |  |
| Primary/None | 3,254 | 1,452 (44.6%) |
| Form 4 | 10,784 | 6,352 (58.9%) |
| Form 6 | 2,233 | 1,702 (76.2%) |
| Post-secondary | 1,411 | 1,145 (81.2%) |
| Employment |  |  |
| Student | 4,963 | 3,479 (70.1%) |
| Employed (formal) | 835 | 585 (70.1%) |
| Employed (informal) | 3,158 | 1,821 (57.7%) |
| None | 8,726 | 4,766 (54.6%) |
| Marital status |  |  |
| Married | 3,559 | 1,511 (42.5%) |
| Single | 13,324 | 8,732 (65.5%) |
| Divorced or widowed | 799 | 408 (51.1%) |

**Figure 1:**
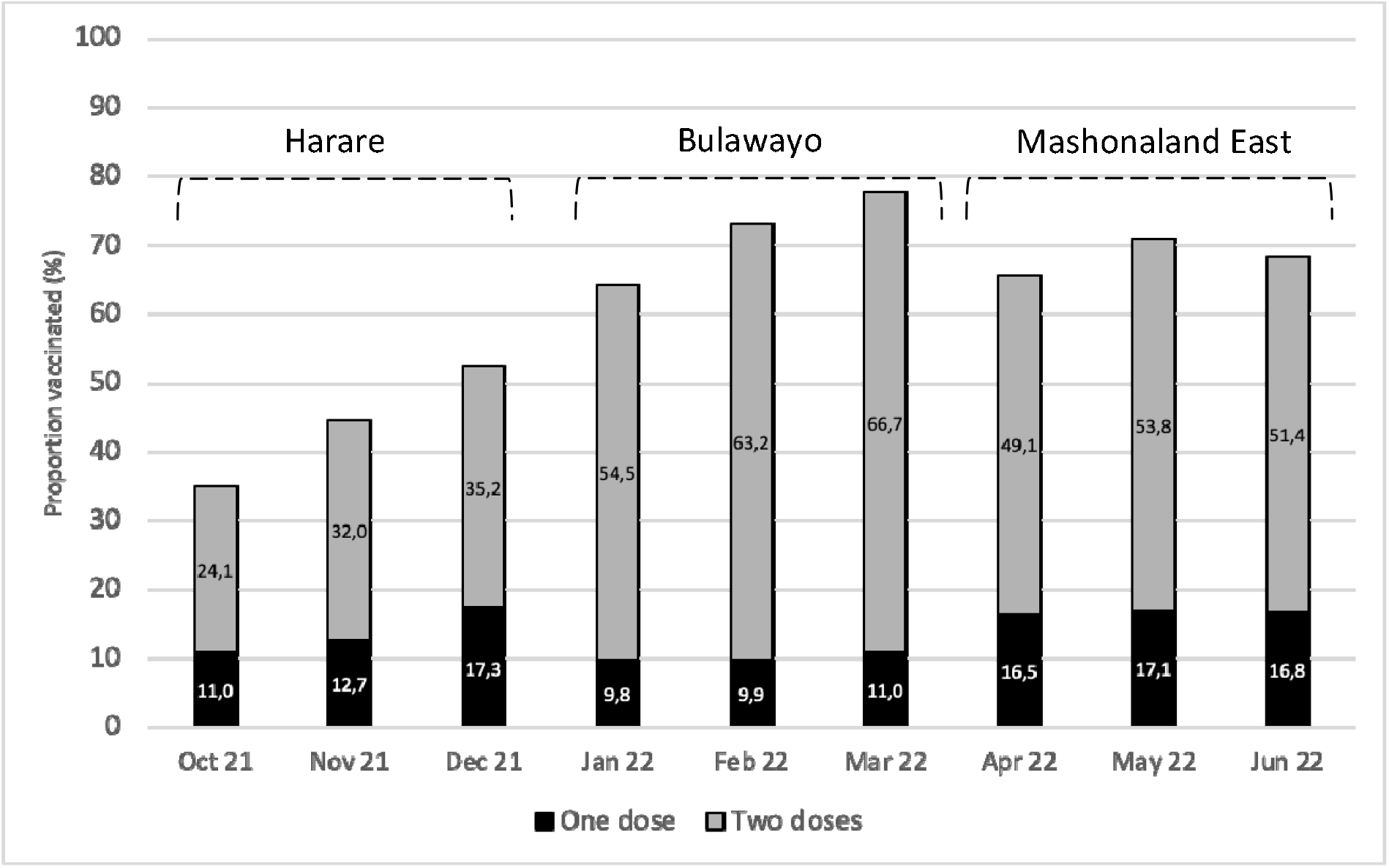
Proportion of young people aged 18-24 who reported being vaccinated by month of the CHIEDZA prevalence survey stratified by province.

The major reasons given for not being vaccinated were lack of time, belief that the vaccine was unsafe, and anxiety about side effects (Figure 2). Men reported a lack of time as the main reason for not getting vaccinated. Women reported concerns related to side effects and safety as reasons for not getting vaccinated more frequently compared to men, especially infertility (10.7%). 755 (4.3%) women in the survey were pregnant at the time of recruitment. Of these women, 297 (39.3%) reported having received at least one dose of a COVID-19 vaccine. Of note, participants frequently reported that they were “afraid to die within 2 years of receiving the vaccine”, 17.7%, 15.6% and 8.2% in Harare, Mashonaland East and Bulawayo respectively. On the other hand, religious belief was a less commonly (< 5.0%) mentioned reason for getting vaccinated. While those living under more deprived socioeconomic circumstances had lower vaccine uptake, reasons for not getting vaccinated were similar across all socioeconomic quintiles (Supplemental Table 1).

**Figure 2:**
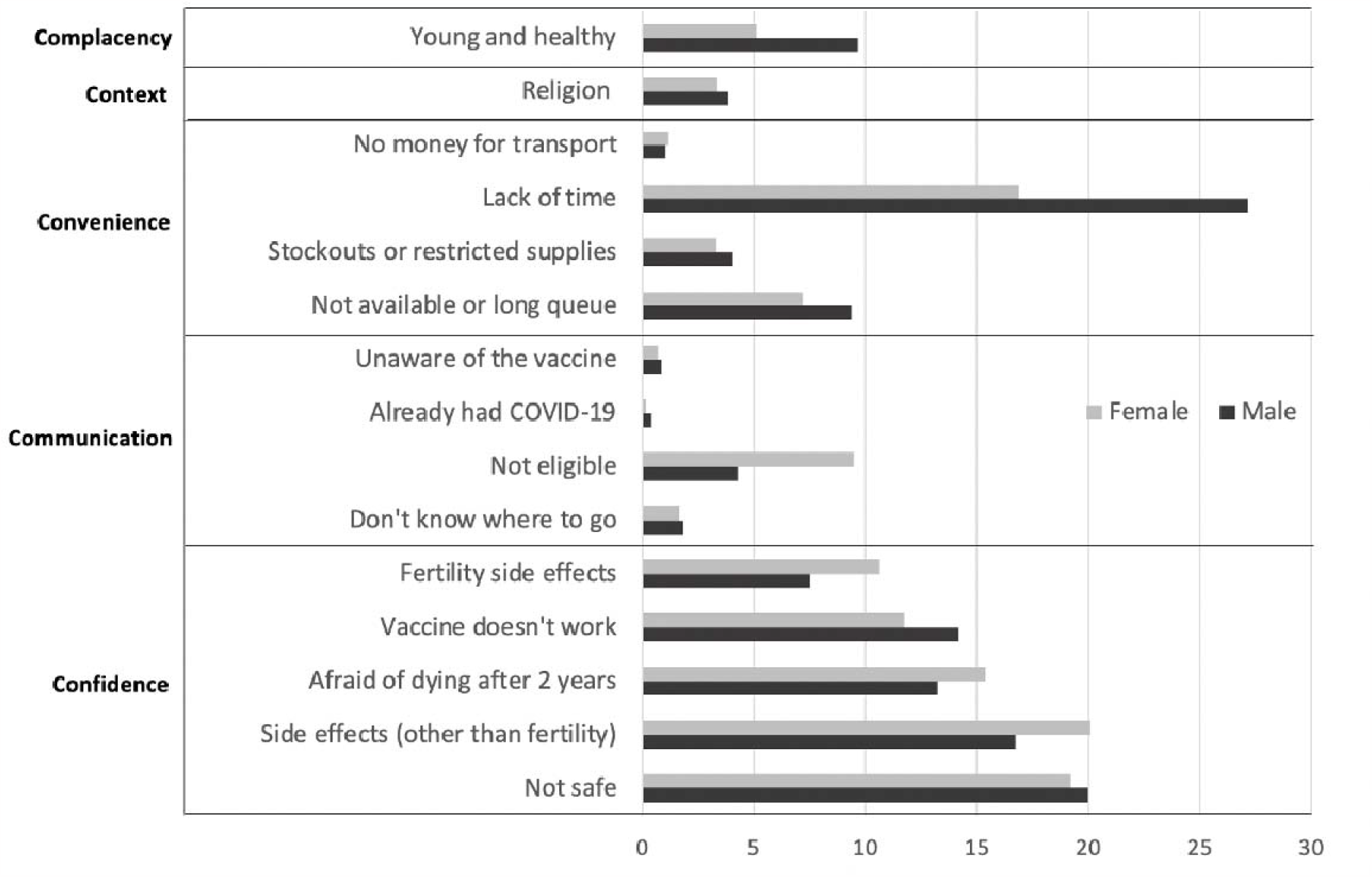
Reasons for not receiving the COVID-19 vaccine stratified by sex and grouped based on the 5C’s of vaccine hesitancy delineated by Razai et al. shown as proportions (%) [31]. Participants could provide multiple reasons and could only answer the question if they were not vaccinated. These reasons were read out or shown to the participant for them to choose from

Univariable analysis (Supplemental Table 2) showed an association between vaccine uptake age, sex, educational attainment and socioeconomic status. The association between vaccine uptake and predictors remained in the multivariable analysis (Table 2). Men (OR 1.69, 95%CI 1.59-1.80), older youth (20-22: OR 1.06, 95%CI 0.98-1.15, >22: OR 1.12, 95%CI 1.04-1.21), those with higher educational attainment (form 4: OR 1.79, 95%CI 1.39-2.30, form 6: OR 3.56, 95%CI 2.72-4.66, post-secondary: OR 4.34, 95%CI 3.27-5.76), and higher socioeconomic status (2^nd^ quintile: OR 1.06, 95%CI 0.96-1.17, 3 ^rd^ quintile: OR 1.12, 95%CI 1.01-1.23, 4 ^th^ quintile: OR 1.17, 95%CI 1.06-1.29, least poor 20%: OR 1.32, 95%CI 1.20-1.47), were more likely to be vaccinated. Overall results and results stratified by province were comparable.

**Table 2:** Multivariable logistic regression analysis of the association between COVID-19 vaccination and sociodemographic variables

|  | Overall<br>OR (95% CI) | Harare<br>OR (95% CI) | Bulawayo<br>OR (95% CI) | Mashonaland<br>East<br>OR (95% CI) |
| --- | --- | --- | --- | --- |
| Sex |  |  |  |  |
| Female | 1 (p < 0.0001) | 1 (p < 0.0001) | 1 (p < 0.0001) | 1 (p < 0.0001) |
| Male | 1.69 (1.59 – 1.80) | 1.45 (1.29 – 1.64) | 1.43 (1.27 – 1.61) | 1.84 (1.63 – 2.08) |
| Age |  |  |  |  |
| < 20 | 1 (p = 0.0123) | 1 (p < 0.0001) | 1 (p = 0.0002) | 1 (p = 0.6537) |
| 20-22 | 1.06 (0.98 – 1.15) | 1.23 (1.06 – 1.42) | 1.24 (1.08 – 1.43) | 0.94 (0.81 – 1.08) |
| > 22 | 1.12 (1.04 – 1.21) | 1.43 (1.25 – 1.63) | 1.31 (1.14 – 1.50) | 0.99 (0.86 – 1.14) |
| Education |  |  |  |  |
| Primary | 1 (p < 0.0001) | 1 (p < 0.0001) | 1 (p < 0.0001) | 1 (p < 0.0001) |
| Form 4 | 1.79 (1.39 – 2.30) | 1.81 (1.15 – 2.86) | 1.83 (1.15 – 2.93) | 1.39 (0.88 – 2.20) |
| Form 6 | 3.56 (2.72 – 4.66) | 4.09 (2.55 – 6.57) | 3.50 (2.13 – 5.75) | 2.84 (1.73 – 4.66) |
| Secondary and above | 4.34 (3.27 – 5.76) | 5.20 (3.19 – 8.47) | 4.44 (2.60 – 7.58) | 3.51 (2.08 – 5.92) |
| Socioeconomic status |  |  |  |  |
| Poorest | 1 (p < 0.0001) | 1 (p < 0.0001) | 1 (p = 0.3776) | 1 (p = 0.0010) |
| 2 | 1.06 (0.96 – 1.17) | 1.26 (1.05 – 1.50) | 1.04 (0.89 – 1.22) | 0.95 (0.81 – 1.13) |
| 3 | 1.12 (1.01 – 1.23) | 1.38 (1.16 – 1.64) | 0.99 (0.78 – 1.25) | 0.99 (0.83 – 1.18) |
| 4 | 1.17 (1.06 – 1.29) | 1.70 (1.43 – 2.03) | 1.10 (0.92 – 1.32) | 1.01 (0.83 – 1.23) |
| Least poor | 1.32 (1.20 – 1.47) | 1.93 (1.61 – 2.31) | 1.09 (0.90 – 1.31) | 1.32 (1.09 – 1.61) |

## Discussion

This study found a COVID-19 vaccine coverage among young people aged 18-24 years of 69.6% in Bulawayo, 68.3% in Mashonaland East, and 42.6% in Harare. Despite not being a high-risk group, the national vaccination campaign reached them effectively. Vaccine uptake was however inequitable. Those who were male, older, more educated, and of higher socioeconomic status were more likely to report COVID-19 vaccination, which is in-line with studies conducted in HICs, though the specific reasons may be different. Education attainment and male sex were the strongest predictors.

The difference in COVID-19 vaccine coverage among youth across the provinces is partly explained by the staggered timing of the survey. The national vaccination campaign started in February 2021 prioritising front-line workers. Vaccination eligiblity was extended to all adults in March/April 2021 and 16-18-year-olds became eligible in November 2021 (one month after study recruitment started in Harare). At the time the survey was completed in each province, the provincial vaccine coverage in Zimbabwe was 25.3% (Harare), 31.7% (Bulawayo), and 38.6% (Mashonaland East) [33]. Importantly, in the context of COVID-19 vaccination, coverage has been defined as the percentage of the total population that is vaccinated and includes children even though they may not be eligible [34]. At the end of the survey, vaccine coverage using the total eligible population (16* years) as denominator was 73.8% nationally, and thus comparable to coverage among youth in Mashonaland East [2].

In the full adjusted model, men had 1.69 times the odds to have received the vaccine compared to women. Other studies have found higher proportions of men compared to women reporting intention to getting the COVID-19 vaccine [35-37]. These studies have also highlighted that the difference in intention to gett vaccinated against COVID-19 between men and women is less about increased health-seeking behaviour in men and more about decreased health-seeking behaviour among women, which is consistent with existing literature on general vaccine hesitancy [38,39]. This difference is likely due to specific gender differences in risk aversion and potential side effects. Reasons for not getting vaccinated in our study were mainly related to confidence, i.e. regarding safety and efficacy of the vaccine, especially among women. In this survey, 20.1% of women who were not vaccinated said they were afraid of side effects in general and 10.7% said they were afraid of infertility specifically. Furthermore, 66/458 (14.4%) of unvaccinated pregnant women reported fertility-related fears as a barrier, despite the WHO having recommended the use of SinoPharm in pregnant women [35-37,40]. This disparity may be due to confusing communication regarding pregnancy and breastfeeding at the start of the vaccination campaign, including information by official sources and on social media [41,42]. The infodemic about vaccines spread through social media has undoubtably played an important role in Zimbabwe given the high proportion of individuals (both men and women) who reported being afraid of dying within 2 years of vaccination. This myth relates to a widely and globally circulated text message meme claiming that French virologist Luc Montagnier had said all vaccinated people will “die within two years” [42].

Fear of side effects was less frequent among unvaccinated men compared to women, and a higher proportion of men did not feel at risk (felt ‘young and healthy’) or said they were too busy for vaccination. This is despite a vaccination campaign that tried to bring vaccines to the population by offering transport incentives such as transport money, decentralising vaccination to polyclinics, and providing vaccine outreach services [16].

Vaccination rates varied by socioeconomic status and particularily educational attainment, pointing towards health inequity. Other studies have reported that poorer and less educated people experience more barriers to vaccination [44,45]. Those with lower educational attainment are less likely to have access to accurate information and the vaccine information itself might be inaccessible in terms of language, content, and format, especially if this is provided in a written format [46]. Public health information may also include jargon which discriminates against those of lower education level. Those of lower socioeconomic status were also more likely to experience adverse events such as a cut in household income, inability to access food, higher disease risk, or loss of work than those in higher quintiles, which affects people’s ability to receive a vaccine [47].

In this study only a small proportion of individuals said they were not vaccinated because of their religious beliefs. This is in contrast to the results from studies conducted in other countries in Africa where religious beliefs were among the most frequent reasons given for vaccine hesitancy [10-12]. In Zimbabwe the Apostolic church, a Pentecostal Christian denomination, has an estimated 3.5 million followers mainly among poorer and rural households. Children of members of the Apostolic church have been found to have low childhood vaccination coverage [48]. Religious affiliation was not collected in the survey, but membership of the Apostolic church is less common in urban and peri-urban settings, which may explain some of our findings. Also, the Ministry of Health and Child Care, supported by UNICEF, actively reached out to churches including the Apostolic Church to support vaccine education and public engagement [49]. The Apostolic Women’s Empowerment Trust started a COVID-19 awareness programmes in 2021 with the aim to address vaccine hesitancy among those of Apostolic faith [50].

Strengths of this study include a large representative sample of young people from three provinces at the time of the COVID-19 vaccine roll-out. Participation in the survey was high and results were comparable across all three provinces. The main limitation of this study is that COVID-19 vaccination status was self-reported and social desirability bias may have resulted in overestimation of vaccination coverage. However, the survey included a range of potentially sensitive questions, on topics including sexual and reproductive health, and substantial efforts were invested in training the survey team. The survey was conducted in urban and peri-urban settings only and vaccination coverage may be very different among young people living in rural areas. There were no questions specifying the type of vaccine received or the date of vaccination. Another limitation was that the timing of the survey relative to the COVID-19 vaccination campaign differed between the three provinces. Finally, while we asked young people to provide reasons for not being vaccinated, more detailed questions on beliefs, myths, and sources of information might have provided better understanding. Further qualitative research investigating the barriers towards vaccination, especially among women, would strongly contribute to the findings reported in this study.

## Conclusion

This study showed inequitable COVID-19 vaccine coverage among young people aged 18-24 years in three provinces in Zimbabwe (Harare, Bulawayo, and Mashonaland East). Vaccination rates were lower among women, people with lower educational attainment and among those living in more deprived socioeconomic circumstances. Fear around death and infertility fueled by myth and misinformation were among the main reasons for vaccine hesitancy. Therefore, we recommend that national vaccination campaigns should include a major focus on health education, especially for women and those with less education, and particularly around infertility and death. Information that is clear and consistent provided by trusted sources is crucial, and in this way, social media can be used in a positive manner to combat misinformation. Furthermore, campaigns should focus on community and religious leaders as they have a strong impact on their respective groups. No single programme is likely to address vaccine hesitancy in this population and thus strategies should address these inequities by understanding concerns and tailoring vaccine campaigns to specific groups instead of a one-size-fits-all approach.

## Data Availability

All data produced in the present study are available upon reasonable request to the authors

## Acknowledgements

The CHIEDZA study is funded by the Wellcome Trust (Senior Fellowship to RAF: 206316/Z/17/Z)

