## Supplemental Tables and Figures for "Sociodemographic inequity in COVID-19 vaccine uptake among youth in Zimbabwe" (from another version of this preprint: version 1, 2023-03-10)

**Supplemental Table 1:** Reason for not getting vaccinated stratified by socioeconomic quintile

|  | Poorest 20% (n=1897) | 2nd quintile (n=1341) | 3rd quintile (n=1399) | 4th quintile (n=1322) | Least poor 20% (n=1062) |
| --- | --- | --- | --- | --- | --- |
| I didn't know that there was a vaccine against COVID-19 | 15 (0.8%) | 9 (0.7%) | 12 (0.9%) | 11 (0.8%) | 5 (0.5%) |
| I think I’m not eligible for the COVID-19 vaccine | 136 (7.2%) | 111 (8.3%) | 127 (9.1%) | 106 (8.0%) | 70 (6.6%) |
| I didn’t know where I can get the COVID-19 vaccine | 22 (1.2%) | 24 (1.8%) | 27 (1.9%) | 25 (1.9%) | 21 (2.0%) |
| I do not have money for transport to get to the vaccination centre | 17 (0.9%) | 22 (1.6%) | 17 (1.2%) | 16 (1.2%) | 7 (0.7%) |
| I do not have time to go and get the vaccine | 328 (17.3%) | 256 (19.1%) | 286 (20.4%) | 280 (21.2%) | 246 (23.2%) |
| I have already had COVID-19, so I don’t need to be vaccinated | 1 (0.1%) | 3 (0.2%) | 1 (0.1%) | 11 (0.8%) | 1 (0.1%) |
| I have heard there are not enough vaccines (stockouts/ restricted supplies) in the country | 62 (3.3%) | 48 (3.6%) | 48 (3.4%) | 48 (3.6%) | 41 (3.9%) |
| I went to the vaccination centre but couldn’t get the vaccine (not available or long queue) | 125 (6.6%) | 111 (8.3%) | 117 (8.4%) | 118 (8.9%) | 77 (7.3%) |
| I am young and healthy, so I don't need a vaccine | 80 (4.2%) | 78 (5.8%) | 96 (6.9%) | 117 (8.9%) | 83 (7.8%) |
| The vaccine might hurt my fertility | 201 (10.6%) | 112 (8.4%) | 151 (10.8%) | 126 (9.5%) | 87 (8.2%) |
| I am worried about side effects (other than fertility) | 377 (19.9%) | 228 (17.0%) | 265 (18.9%) | 254 (19.2%) | 207 (19.5%) |
| I don’t think the vaccine is safe | 362 (19.1%) | 223 (16.6%) | 288 (20.6%) | 279 (21.1%) | 206 (19.4%) |
| I don’t believe the vaccine works | 207 (10.9%) | 155 (11.6%) | 176 (12.6%) | 194 (14.7%) | 139 (13.1%) |
| My religion does not allow me to have the COVID-19 vaccine | 75 (4.0%) | 43 (3.2%) | 55 (3.9%) | 44 (3.3%) | 28 (2.6%) |
| Afraid of dying after 2 years | 300 (15.8%) | 190 (14.2%) | 216 (15.4%) | 196 (14.8%) | 125 (11.8%) |

**Supplemental Table 2:** Univariable logistic regression analysis of the association between COVID-19 vaccination and sociodemographic variables

|  | **Overall**  **OR (95%CI)** | **Harare**  **OR (95%CI)** | **Bulawayo**  **OR (95%CI)** | **Mashonaland East**  **OR (95%CI)** |
| --- | --- | --- | --- | --- |
| Sex |  |  |  |  |
| Female | 1 (base) | 1 (base) | 1 (base) | 1 (base) |
| Male | 1.83 (1.72 – 1.95) | 1.68 (1.51 – 1.88) | 1.52 (1.36 – 1.70) | 2.05 (1.82 – 2.30) |
| Age |  |  |  |  |
| < 20 | 1 (base) | 1 (base) | 1 (base) | 1 (base) |
| 20-22 | 1.18 (1.10 – 1.28) | 1.38 (1.21 – 1.58) | 1.36 (1.19 – 1.56) | 1.06 (0.92 – 1.21) |
| > 22 | 1.22 (1.14 – 1.31) | 1.57 (1.39 – 1.78) | 1.44 (1.26 – 1.63) | 1.05 (0.93 – 1.20) |
| Education |  |  |  |  |
| Primary | 1 (base) | 1 (base) | 1 (base) | 1 (base) |
| Form 4 | 1.78 (1.39 – 2.28) | 1.79 (1.14 – 2.79) | 1.89 (1.19 – 3.00) | 1.44 (0.92 – 2.26) |
| Form 6 | 3.80 (2.92 – 4.95) | 4.54 (2.85 – 7.23) | 3.74 (2.29 – 6.11) | 3.17 (1.96 – 5.14) |
| Tertiary | 4.71 (3.57 – 6.21) | 6.15 (3.82 – 9.89) | 4.94 (2.92 – 8.35) | 3.98 (2.39 – 6.61) |
| Socioeconomic status |  |  |  |  |
| Poorest | 1 (base) | 1 (base) | 1 (base) | 1 (base) |
| 2 | 1.21 (1.10 – 1.32) | 1.38 (1.16 – 1.64) | 1.19 (1.02 – 1.39) | 1.01 (0.86 – 1.19) |
| 3 | 1.38 (1.26 – 1.51) | 1.72 (1.46 - 2.03) | 1.20 (0.95 – 1.50) | 1.21 (1.02 – 1.42) |
| 4 | 1.54 (1.41 – 1.69) | 2.28 (1.94 – 2.69) | 1.36 (1.12 – 1.59) | 1.39 (1.15 – 1.67) |
| Least poor | 1.98 (1.80 – 2.18) | 2.86 (2.42 – 3.38) | 1.51 (1.27 – 1.80) | 2.02 (1.69 – 2.43) |

Adjusted by month of recruitment, OR=odds ratio, 95%CI=95% confindence interval
